# Cocoa flavanols improve peakVO_2_ and exercise capacity in a randomized double blinded clinical trial in healthy elderly

**DOI:** 10.1101/2023.04.25.23289076

**Authors:** Michael Gröne, Dragos Duse, Nicolas Kramser, Niklas Ophoff, Hendrik Schweers, Fabian Voß, Christine Quast, Roberto Sansone, Christian Heiss, Christian Jung, Malte Kelm, Ralf Erkens

## Abstract

**Background:** Loss of functional capacity is one of the hallmarks in cardiovascular aging. Cocoa flavanols (CF) exert favorable effects on endothelial function, blood pressure, and inflammation. These cardiovascular health markers worsen with increasing age and limit functional exercise capacity.

**Aim:** To investigate the effect of CF on cardiorespiratory-fitness in healthy elderly.

**Methods:** In a randomized, double-masked, placebo-controlled, parallel-group dietary intervention trial, 68 healthy elderly (55-79 years, 28 female) received either 500 mg of CF or a nutrient-matched control capsule twice a day for 30 days. Primary endpoint was defined as peak oxygen consumption (VO_2_) in a cardiopulmonary exercise test (CPET). Secondary endpoints were oxygen pulse (VO_2_/heart rate (HR)), resting blood pressure (BP), and resting vascular function.

**Results:** After 30 days of CF intake peakVO_2_ increased by 190 ml/min (95% CI 1-371 ml/min) and peakVO_2_/kg by 2.5 ml/(min*kg) (95% CI 0.30-4.2 ml/(min*kg)). O_2_-pulse increased by 1.7 ml (95% CI 0.29-3.2 ml) and max exercise capacity by 9.6 W (95% CI 2.1-17.7 W). CF decreased resting systolic and diastolic BP by 5.4 mmHg (95% CI -10.7 - -0.1 mmHg) and 2.9 mmHg (95% CI (-) 5.5-(-) 0.4 mmHg), respectively. Flow-mediated vasodilation (FMD) increased by an absolute 1.3% (95% CI 0.76-1.79 %) in the CF group. Indexes of pulmonary function were not affected. No changes for primary and secondary endpoints were detected in control.

**Conclusion:** CF substantially improve markers of cardiorespiratory fitness in healthy elderly humans highlighting their potential to preserve cardiovascular health with increasing age.

## Introduction

Loss of functional capacity is a hallmark of cardiovascular aging. Age is one of the major risk factors for development of cardiovascular disease (CVD) (1, 2). The aging heart is characterized by increasing mass-to-volume ratio, reduced stroke volume and reduced maximum heart rate resulting in reduced cardiac output, which diminishes cardiac reserve capacity and functional status (3). Reduced cardiac regeneration capacity, oxidative stress and chronic low-grade inflammation are key factors for the development of age associated cardiac disease (4). Additionally, aging is associated with endothelial dysfunction and arterial remodeling culminating in luminal dilation, intimal thickening (5, 6), which overall predispose for the development of vascular stiffness.

Cardiovascular fitness as a strong predictor of mortality (7), is of socioeconomic importance in an aging population worldwide, where cost for care and nursing are growing (8). Cardiopulmonary exercise testing (CPET) is the standard non-invasive method for the quantification of exercise capacity and discrimination of underlying reasons for functional limitation (9). The ability of CPET to assess both peak and submaximal exercise response in a ramp protocol makes it a useful tool in the quantification of capability in daily activities. CPET is a valuable method to estimate prognosis in patients with heart failure and CAD (10). Peak oxygen consumption (peakVO_2_) has been independently associated with all-cause mortality, death from heart failure, sudden cardiac death, and functional deterioration in heart failure and hypertrophic cardiomyopathy (3, 10). High levels of cardiorespiratory fitness are associated with the lowest risk-adjusted all-cause mortality (7).

CF exert favorable effects on several cardiovascular health markers including low-grade inflammation, oxidative stress (11), endothelial function (12), and blood pressure (13). Improved endothelial function and reduced vascular stiffness and normotensive blood pressure positively affect ventricular-arterial-coupling (14) leading to improved cardiac output and supposedly better exercise performance. Whether CF have a positive impact on peakVO2 and exercise capacity in the older adults remains unknown. The aim of the current study was to investigate the effect of dietary CF intake on cardiorespiratory fitness promoting functional status in the healthy elderly. For this, capsules containing a high-flavanol cocoa extract were given in a randomized, placebo-controlled, double-blinded trial to healthy elderly humans following structured exercise testing.

## Methods

### Study design

In a randomized, double-masked, placebo-controlled, parallel-group dietary intervention trial, 68 healthy elderly humans (55-79 years, 28 female) were recruited to undergo CPET, vascular function assessment and biomarker testing. Inclusion criteria was healthy elderly participants over 55 years of age and without diagnoses of cardiovascular disease. In addition to past medical history, heart rate, electrocardiogram, physical examination and laboratory parameters validated healthy status in all participants (Table 1). Exclusion criteria included age under 55 or over 80 years, non-cardiac limitations of exercise capacity such as orthopedic or neurological disorders, BMI >35 kg/m^2^, active smoking, uncontrolled blood pressure, diabetes, chronic obstructive pulmonary disease, atrial fibrillation, cardiomyopathy, coronary heart disease, valvular heart disease, cardiac pacemaker, acute respiratory tract infections, regular intake of any kind of medication and missing ability or missing cooperation to participate in the study.

**Table 1:** Baseline characteristics of the study cohort. There were no significant differences between groups at baseline, values are reported as means ± standard deviation (SD). BSA = body surface area; HbA1C = glycated hemoglobin; CRP = C-reactive protein; Hb = hemoglobin.

| Baseline Characteristics | Control (n=33) | Cocoa flavanols (n=35) | p-value |
| --- | --- | --- | --- |
| Age (y) | 64.5 ± 6.6 | 64.3 ± 5.4 | 0.8884 |
| Sex |  |  |  |
| Male | 17 | 23 |  |
| Female | 16 | 12 |  |
| Height (cm) | 174 ± 8.4 | 177 ± 8.9 | 0.0996 |
| Weight (kg) | 80.2 ± 13.0 | 82.5 ± 14.1 | 0.4839 |
| BSA (m<sup>2</sup>) | 1.99 ± 0.20 | 1.98 ± 0.19 | 0.8296 |
| *Blood Pressure (mmHg) - Baseline* |  |  |  |
| Systolic | 139.5 ± 16.8 | 137.3 ± 14.6 | 0.3421 |
| Diastolic | 81.0 ± 9.5 | 80.3 ± 8.5 | 0.7555 |
| Heart Rate (b/min) - Baseline | 69.6 ± 12.7 | 67.6 ± 10.4 | 0.4654 |
| *Cholesterol (mg/dl)* |  |  |  |
| Total | 208 ± 41.6 | 212 ± 37.7 | 0.6815 |
| Low-density | 126 ± 36.0 | 133 ± 33.9 | 0.4125 |
| High-density | 65 ± 17.8 | 64 ± 22.0 | 0.8500 |
| Triglycerides (mg/dl) | 170 ± 154.5 | 163 ± 93.6 | 0.8034 |
| Creatinine (mg/dl) | 0.84 ± 0.15 | 0.89 ± 0.19 | 0.2194 |
| HbA1C (%) | 5.48 ± 0.47 | 5.43 ± 0.32 | 0.6214 |
| CRP (mg/dl) | 0.25 ± 0.25 | 0.16 ± 0.13 | 0.0594 |
| Hb (g/dl) | 14.01 ± 0.76 | 13.87 ± 2.37 | 0.7367 |
| White blood cells (*1000/μL) | 6.78 ± 1.25 | 6.64 ± 1.31 | 0.6442 |

After baseline assessment, participants were randomized to receive either 500 mg of high-flavanol cocoa extract (CF) or a nutrient-matched control capsule twice a day for 30 days. The total amount of flavanols in mg represents the sum of all monomeric flavanols and oligomers, as published by us (15). The predominant monomeric flavanol in our capsule was (-) –epicatechin (Table S1). Capsule format was utilized to simulate effects of a flavanol-rich diet. Placebo capsules were filled with microcrystalline cellulose and matched for theobromine and caffeine content. Thus, flavanols and (-)-epicatechin were absent in placebo capsules. Overall appearance and weight of placebo capsule were not distinguishable from those of active capsules. After 30 days, the same testing protocol was applied (see Figure 1 A, Study protocol). Written informed consent was obtained from each participant. The study protocol conforms to the ethical guidelines of the 1975 Declaration of Helsinki as reflected in a priori approval the ethics committee of the Heinrich Heine University Düsseldorf (Approval Number R5761R). Registered on clinicaltrials.gov (NCT 05782309). Unblinding process was performed by a pre-determined protocol.

**Fig. 1:Study protoco.**
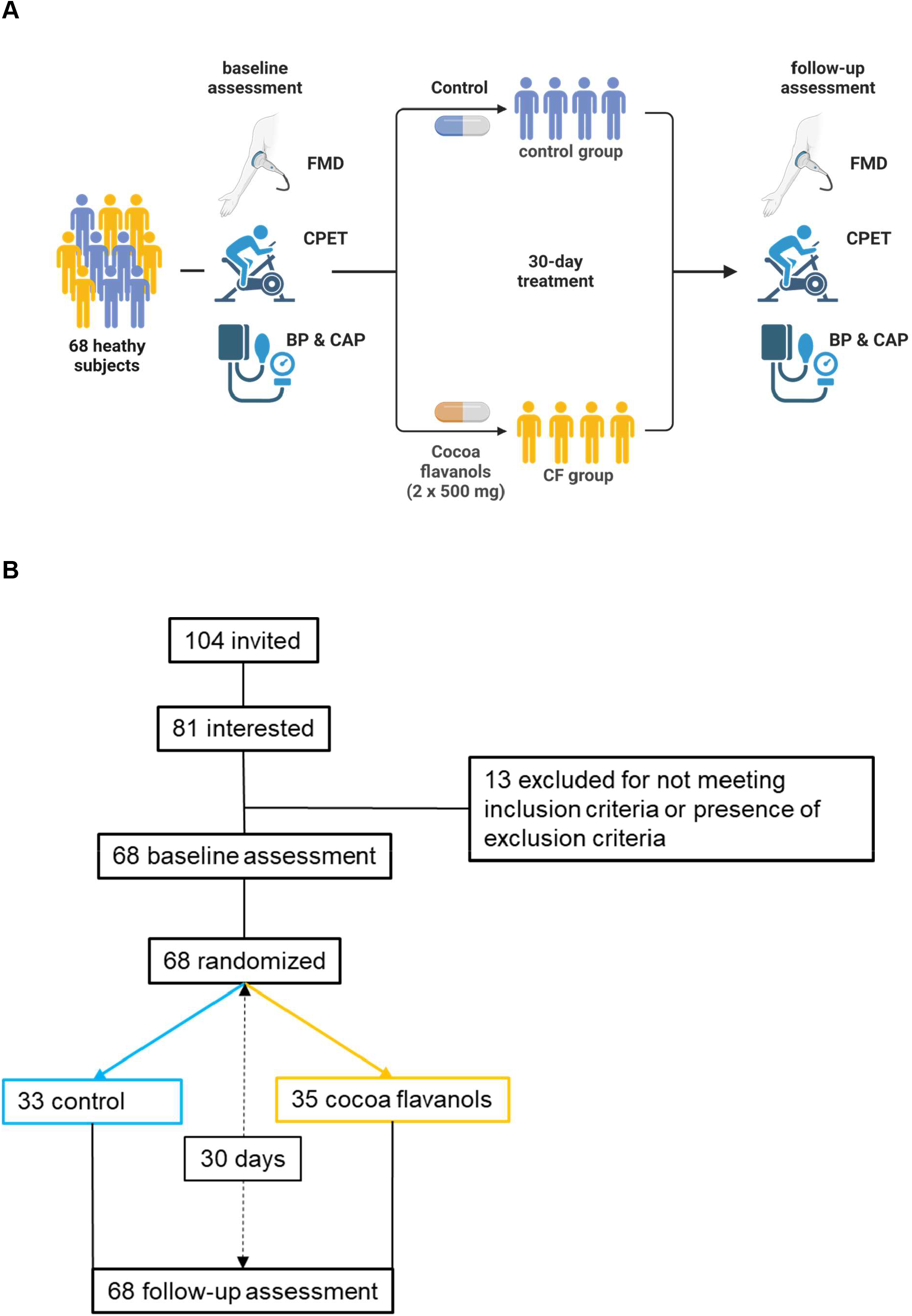
**Fig 1A:** l68 healthy elderly adults (aged 55 to 79 years) received either 500 mg of Cocoa flavanols (CF) capsule twice daily or a nutrient-matched control capsule for 30 days in a randomized, double-masked, placebo-controlled design. Cardiopulmonary exercise testing (CPET), flow-mediated dilatation (FMD), central aortic pressure (CAP) and blood pressure (BP) measurements were performed at baseline and after 30 days. Primary endpoint was exercise capacity and peakVO_2_. Secondary endpoints included CAP, BP, and resting vascular function, measured as FMD. **Fig. 1 B:** CONSORT flow diagram.

### CPET

CPET was conducted using a standardized bicycle ergometer test protocol at baseline and after 30 days. The test was performed with an upright cycle ergometer at the same time for all patients in the early afternoon. The testing protocol was an incremental ramp protocol with increases in work rate of 25 watts every 2 minutes. Site technicians were trained and certified for the protocol and followed a detailed CPET manual. PeakVO2 was assessed as the primary endpoint. Also, the following prespecified exploratory cardiovascular parameters were recorded: VCO2, peak minute ventilation (VE), peak RER, peak heart rate, peak exercise capacity, peak oxygen pulse ((VO_2_/HR) and VO_2_ at ventilatory threshold. A trained Sports Cardiologist in a double-blinded approach analyzed the CPET data.

### FMD

FMD was measured as previously described (16, 17) on the participant’s right arm. Vessel diameter and flow velocity of the radial artery (RA) were measured using a 12 MHz transducer (Vivid I, GE) and automatic edge-detection software (Brachial Analyzer, Medical Imaging Applications, Iowa City, Iowa) yielding standard deviations of mean differences between repeated measurements of less than an absolute 1.0 %. Reactive hyperemia was induced by 5 min of lower arm occlusion with a sphygmomanometer cuff inflated to 200 mmHg. Immediately after cuff deflation, and 20, 40, 60, and 80 sec later, RA diameter was assessed and FMD was calculated as maximal relative diameter gain relative to baseline.

### Blood pressure and central aortic pressure

Peripheral blood pressure was measured with an automated medical device (boso medicus, BOSCH + SOHN GmbH u. Co. KG) at both participant’s left and right brachial artery to calculate the arithmetical mean, which was the used for analyses.

Central aortic pressure was assessed by noninvasive measurement of the central aortic pressure waveform using Sphygmocor® platform (Atcor medical, Sydney, Australia). Calculation of central aortic pressures is calculated via an integrated algorithm after entering individual patient characteristics and peripheral blood pressure (15).

### Statistical analysis

Data are presented as means ± standard deviation (SD). Statistical analyses were performed by using Fisher’s exact test (case controls), and unpaired t-tests for detecting differences between the groups. Repeated measurements were calculated as ANCOVA with baseline values as covariates with the Bonferroni post hoc test. Significance was assumed if p was < 0.05. Analyses were performed by GraphPad Prism version 9 and SPSS statistics, version 24 (IBM, Armonk, USA). The sample size was calculated prior by using G-Power V3.1. (Heinrich Heine University of Duesseldorf. Significance was assumed if p was < 0.05. Analyses were performed using GraphPad Prism version 9 (GraphPad Software, Inc.) and SPSS statistics, version 24 (IBM, Armonk, USA).

## Results

### Study population

From April 2018 to November 2021, after screening of 104 possible participants, 68 elderly adults (>55 years) fulfilled inclusion criteria and were defined as healthy. None of the participants had previously diagnosed cardiovascular diseases, nor took any kind of medication. Participants were randomized to receive either a CF capsule twice daily or a nutrient matched placebo control capsule. One participant initially randomized to control declined participation after baseline examination and was excluded from the study. Baseline characteristics are presented in Table 1. Baseline exercise parameters were similar for both groups (Table 2). Baseline measures of FMD, peripheral blood pressure and central aortic pressure are presented in Table 3 and were not different between groups.

**Table 2:** Effects of dietary CFs on measures of exercise capacity during CPET. peakVO_2_, peakVO_2_/kg, O2-pulse, and exercise capacity significantly improved with CFs; VE increased in both groups. BL = baseline; FU = follow up; HR = heart rate; RER = respiratory exchange ratio; VE = minute ventilation; peakVO_2_= maximum oxygen uptake, peakVO_2_/kg = maximum oxygen uptake per kg of bodyweight, O_2_-Pulse = oxygen uptake per heartbeat; VT = tidal volume W = Power in Watts; * p < 0.05; repeated measurements 2-way-(time × intervention) ANCOVA with baseline values as covariates with Bonferroni’s post hoc test.

| Measures of exercise capacity during CPET |  |  |  |  |  |  |
| --- | --- | --- | --- | --- | --- | --- |
| Intervention group | Control (n=33) |  |  | Cocoa flavanols (n=35) |  |  |
|  | BL | FU | p | BL | FU | p |
| VO<sub>2</sub>peak (l/min) | 1.88 ± 0.60 | 1.93 ± 0.62 | 0,7742 | 1.83 ± 0.60 | 2.02 ± 0.60 | **\*0,0135** |
| VO<sub>2</sub>peak/kg (ml/min/kg) | 22.40 ± 6.74 | 23.40 ± 6.94 | 0.4790 | 22.74 ± 6.88 | 25.25 ± 6.78 | **\*0.0124** |
| VO<sub>2</sub> at VT1 (l/min) | 1.33 ± 0.48 | 1.41 ± 0.48 | 0,7150 | 1.37 ± 0.45 | 1.36 ± 0.42 | 0.9396 |
| VO<sub>2</sub> at VT1/VO<sub>2</sub>peak [%] | 70.1 % | 70.4 % | --- | 74.9 % | 67.7 % | --- |
| O<sub>2</sub>-pulse (VO<sub>2</sub>/HR) (ml) | 13.29 ± 4.55 | 14.22 ± 4.55 | 0.3316 | 13.37 ± 4.78 | 15.11 ± 3.95 | **\*0.0175** |
| Exercise capacity (W) | 150.5 ± 49.04 | 151.3 ± 51.94 | 0,9667 | 151.5 ± 45.6 | 161.1 ± 53.8 | **\*0,0472** |
| HR max (beats/min) | 140.6 ± 20.4 | 140.7 ± 22.1 | 0.9929 | 136.0 ± 19.7 | 131.6 ± 17.7 | 0.3611 |
| RER max | 1.110 ± 0.09 | 1.091 ± 0.08 | 0,3310 | 1.083 ± 0.08 | 1.075 ± 0.06 | 0.9992 |
| VE max (l/min) | 57.55 ± 21.0 | 64.03 ± 26.4 | **\*0,0229** | 55.26 ± 17.1 | 63.69 ± 22.3 | **\*0,0025** |
| VT max (l) | 2.058 ± 0.68 | 2.153 ± 0.65 | 0,3722 | 2.025 ± 0.59 | 2.144 ± 0.63 | 0.2184 |

**Table 3:**
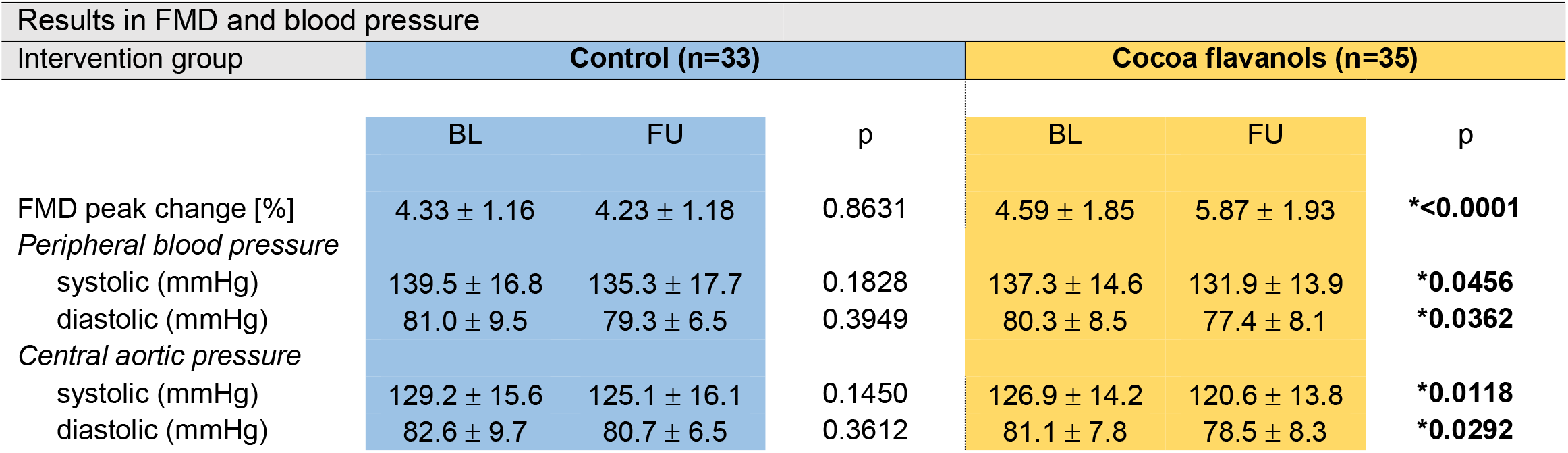
Effects of dietary CFs on vascular function. FMD increased with CFs. Peripheral blood pressure and central aortic pressure decreased with CFs at follow up. No changes were seen in the control group. BL = baseline; FU = follow up FMD = flow-mediated-dilation; * p < 0.05; repeated measurements 2-way-(time × intervention) ANCOVA with baseline values as covariates with Bonferroni’s post hoc test.

### CPET parameters

CF improved peak oxygen uptake (peakVO_2_) by 190 ml (95% CI 1 – 371 ml/min) and peakVO_2_ per kg of bodyweight (VO_2_peak/kg) by 2.51 ml/(min*kg) (95% CI 0.30 – 4.27 ml/(min*kg). O_2_-pulse increased by 1.74 ml (95% CI 0.29-3.18 ml) and maximum exercise capacity by 9.6 W (95% CI 2.1-17.7 W). No significant changes were seen for these parameters in the control group (Figure 2). Maximum minute ventilation (VE) increased in CF and control group. No changes were observed in maximum heart rate (HR max) and RER max. VO_2_ at estimated ventilatory threshold (VO_2_ at VT1) remained unchanged in both groups after 30 days (Table 2). Taken together these data imply that CF increase cardiorespiratory capacity in healthy elderly men and women.

**Fig. 2:**
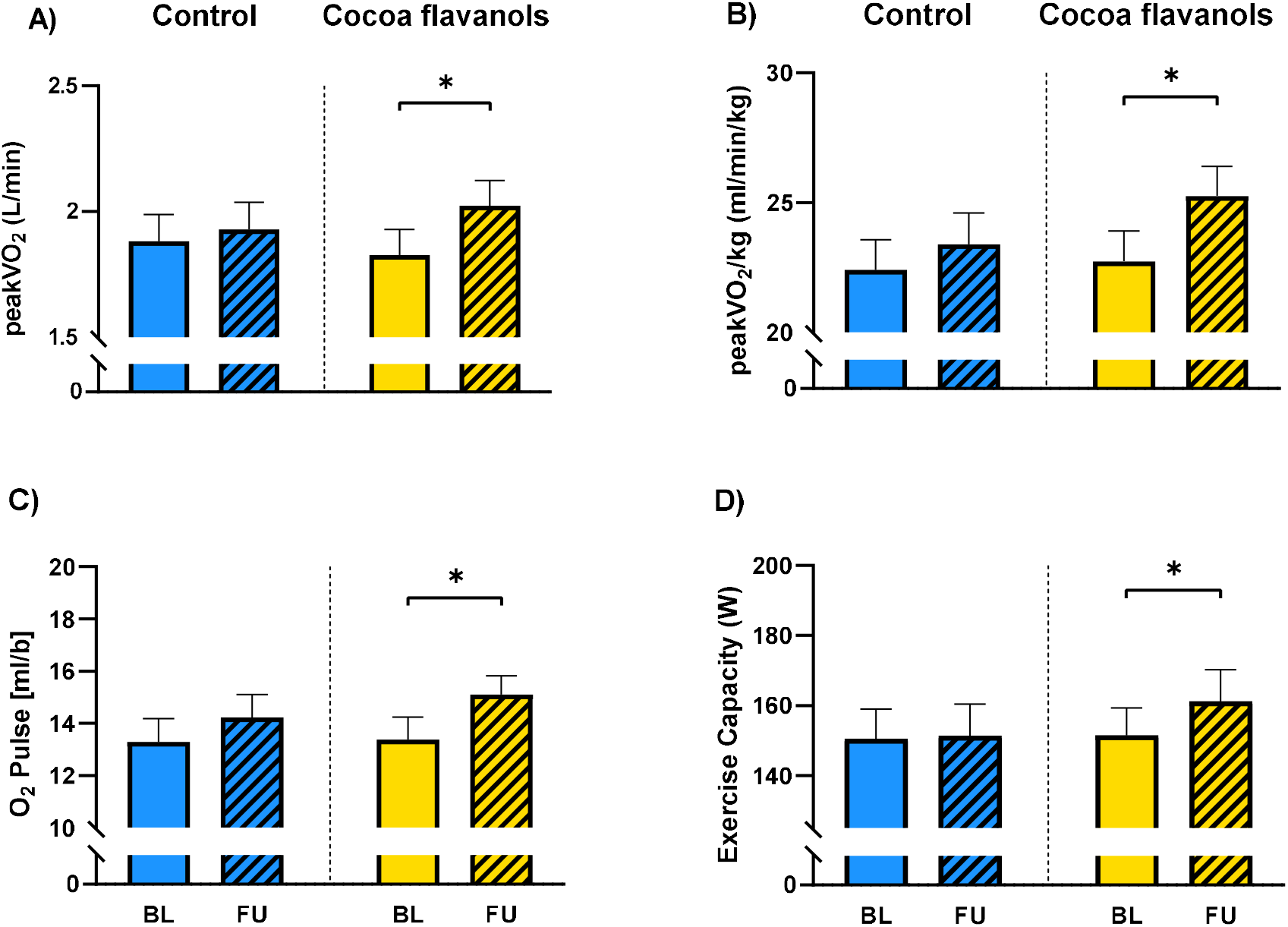
Dietary CFs increase maximum oxygen uptake and exercise capacity. Significant increase in **(A)** VO_2_peak, **(B)** VO_2_/kg, **(C)** O_2_ Pulse, and **(D)** exercise capacity at maximum workload after 30 days of CF intake compared to baseline. No changes were observed with control. * p < 0.05; repeated measurements 2-way-(time × intervention) ANCOVA with baseline values as covariates with Bonferroni’s post hoc test. BL = baseline; FU = follow-up; peakVO2= maximum oxygen uptake, peakVO_2_/kg = maximum oxygen uptake per kg of bodyweight, O_2_-Pulse = oxygen uptake per heartbeat; statistical differences were calculated with paired t-test within group.

### Blood pressure, biomarker and vascular function parameters

Resting systolic and diastolic BP decreased by 5.4 mmHg (95% CI (-) 10.7-(-) 0.1 mmHg) and 2.9 mmHg (95% CI (-) 5.5-(-) 0.4 mmHg), respectively in the CF group. Also, central systolic and diastolic aortic pressure decreased in the CF group by -6.3 mmHg (95% CI (-) 11.1 – (-) 1.5 mmHg) and 2.6 mmHg (95% CI (-) 5.1 – (-) 0.2 mmHg), respectively (Figure 3). FMD increased by an absolute 1.28% (95% CI 0.76-1.79 %) in the CF group. No changes were documented in the control group (Figure 4). In laboratory analyses levels of NT-proBNP were within the normal range for the assay utilized (ULN 125 ng/L) and remained unchanged in both groups.

**Fig. 3:**
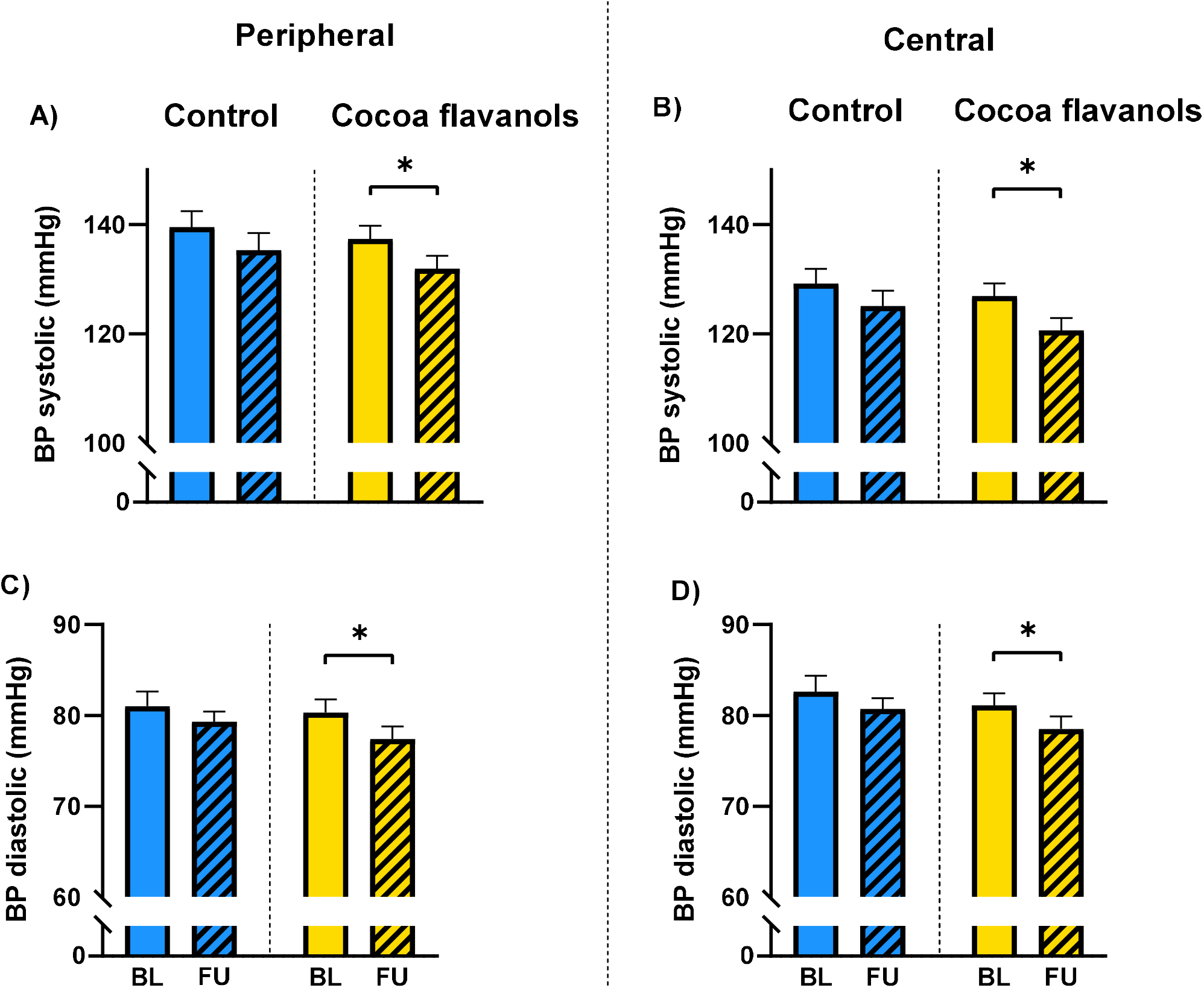
Dietary CFs decrease blood pressure. Significant decrease in peripheral blood pressure **(A & C)** and central aortic pressure **(B & D)** after 30 days of CF intake. No changes occurred with control. * p < 0.05; repeated measurements 2-way-(time × intervention) ANCOVA with baseline values as covariates with Bonferroni’s post hoc test. AP = aortic pressure; BP = blood pressure; BL = baseline; FU = follow up.

**Fig. 4:**
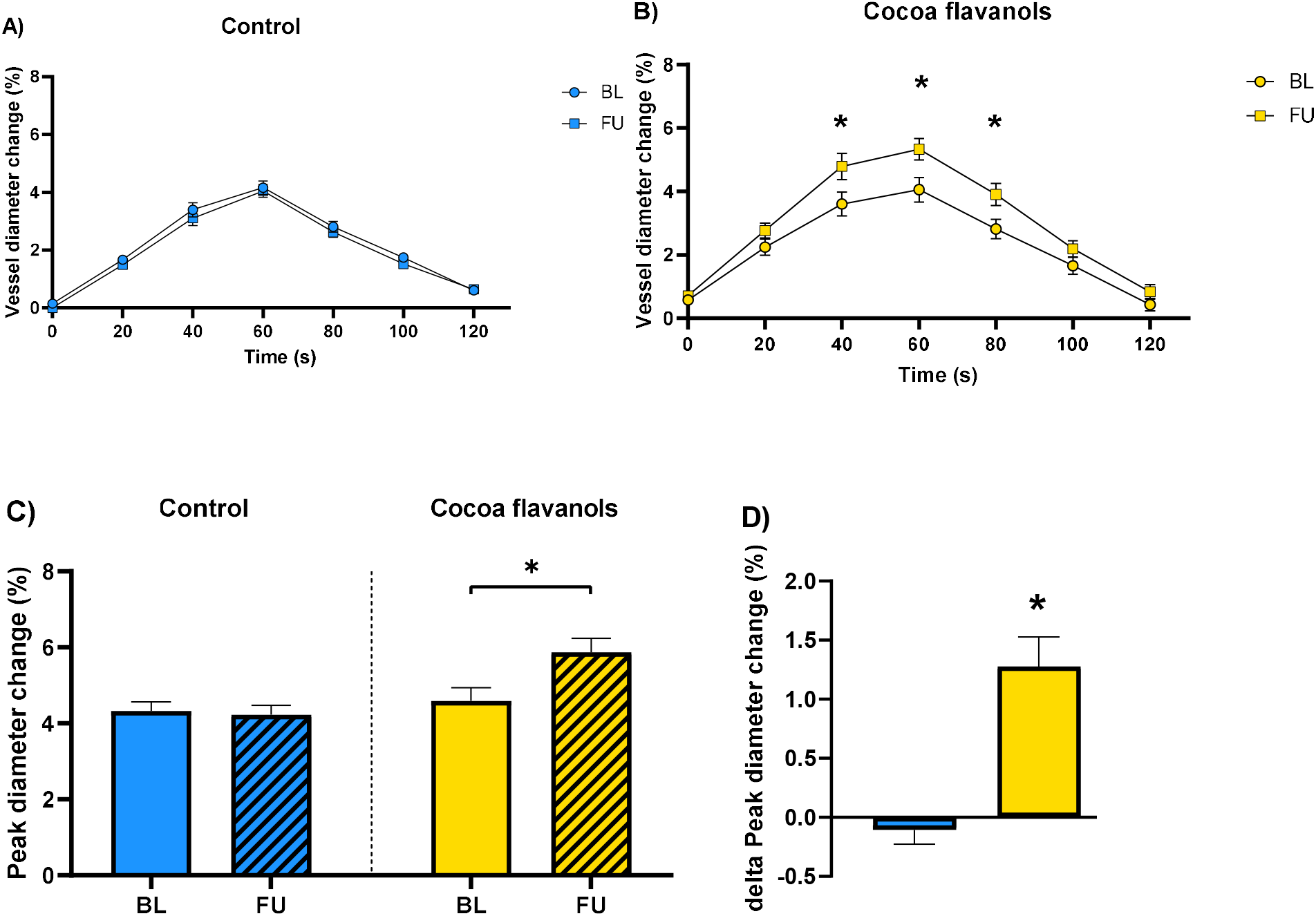
Dietary CFs improve endothelial function. (A) Unchanged FMD with control **(B)** significant increase at FU in FMD at 40s, 60s and at 80s after CF intake compared to BL; **(C)** Significantly higher Peak inner diameter change at FU vs BL with CFs, **(D)** Increased delta in peak diameter change from BL to FU with CFs but not with control; * p < 0.05; repeated measurements 2-way-(time × intervention) ANCOVA with baseline values as covariates with Bonferroni’s post hoc test. BL = baseline; FU = follow up. FMD = flow-mediated-dilation.

## Discussion

The present results clearly demonstrate that dietary CF intake improves exercise capacity in healthy elderly humans. Improvement of cardiorespiratory fitness through dietary intervention may contribute to the maintenance of health in older individuals. The increase in exercise capacity was mainly driven by improvement of cardiac performance while pulmonary indices were not altered. The chosen amount of CFs enhanced endothelial function and lowered arterial blood pressure, both of which may have contributed to improvement in cardiac exercise capacity with significantly increased oxygen carrying capacity per heartbeat.

### Exercise capacity in healthy elderly humans

According to the World Health Organization health is considered a “state of complete physical, mental and social well-being and not merely the absence of disease” (18). Participants in this study were screened prior to randomization to ensure the absence of manifest cardiovascular diseases. With increasing age large artery stiffness develops inducing mild increases in central arterial blood pressure (19). In our study population, elderly participants showed slightly elevated systolic blood pressure with an average of 139 and 137 mmHg. Also, LDL-cholesterol levels were mildly elevated. According to current guidelines lifestyle interventions are recommended as first line option to modify these parameters (20-22). Healthy aging comprises the primary prevention of functional limitations, occult and manifest disease with increasing age. Blood pressure, pulse wave velocity and FMD have been proposed to monitor the transition of healthy to dysfunctional arteries with increasing age (23). Young adults under 25 years of age are generally suspected healthy. After the age of 45 years subclinical disease starts building up depending on previous lifestyle behavior and risk factors. Beyond 65 years of age CVD starts manifesting and eliciting events (24). Age per se is considered an independent risk factor for CVD and mortality (25, 26). CPET is a valuable tool to assess cardiac, circulatory, and pulmonary function and health. It is a highly reliable and reproducible method for measuring peakVO_2_ and to evaluate the functional capacity in healthy and diseased humans. PeakVO_2_ is also a strong predictor of long-term all-cause, CV- and cancer mortality (7, 27). Limitations to cardiopulmonary fitness can often be assigned to one of the primary components: skeletal muscle oxygen extraction, pulmonary oxygen uptake, blood oxygen transport capacity and cardiac output for oxygen distribution. Cardiac performance in this regard is also modulated by preload and afterload. Cardiac afterload is amongst others determined by blood pressure and endothelial function, alterations of both have been associated with the cardiovascular aging process (15, 23). The aging heart itself is less well understood, but comprises a gradual deterioration of functional and structural characteristics including increased mass-to-volume-ratio and a decline in diastolic function (3, 28). Reduced ß-adrenergic response and consecutively reduced maximum heart rate are prevalent in aging hearts (29). Improving cardiorespiratory fitness through progressive exercise training is considered an effective method to reduce cardiovascular functional deterioration in sedentary middle-aged individuals (30). Especially, age-related decline in cardiac functional parameters can be positively modified (31) with aerobic interval training, while the effect of dietary interventions with CFs on exercise capacity in the elderly healthy population has not been well characterized.

### Cocoa flavanols improve exercise capacity in healthy elderly humans

After a period of 30-days dietary intake CF, CF increased peakVO_2_, peakVO_2_/kg, O_2_-pulse, and exercise capacity suggesting beneficial effects on cardiovascular fitness. Indeed, the increase in peakVO_2_ of 190 ml or 2.5 ml/kg of body weight that was achieved with dietary intake is comparable to the effects in structured exercise programs (32, 33). In older adults over 70 years of age, a supervised exercise training with high-intensity intervals improved peakVO_2_ (34). With increasing age applicability of physical exercise programs may be limited due to orthopedic diseases and thus dietary interventions may offer complementary tools to maintain cardiovascular health and fitness.

Maximum heart rate was unaffected in our study groups. An increased O2-pulse suggests an improvement in stroke volume as a potential primary mechanism for improved oxygen transport. Stroke volume is amongst others dependent on the afterload. Determinants of left ventricular afterload are BP, CAP and aortic stiffness (23, 35), all of them have been positively modulated through CFs. In our study population a reduction in systolic BP by 5.4 mmHg and CAP by 6.3 mmHg was achieved. Thus an improved afterload and arterio-ventricular coupling can contributed to the increase in stroke volume. Decreased afterload, in the long term, prevents ventricular remodeling and hypertrophy (36). To support this process through a different mechanism, CFs have been linked to decreased inflammatory response (37), which further reduces progression of left ventricular remodeling, hypertrophy and fibrosis (26, 38, 39).

Changes in markers of pulmonary function were not linked to increased exercise capacity or peakVO2 in our study. Exercise induced increases in VT were not different between the groups and VE increased in both CF and control group to a similar extent.

### Effectiveness of CFs to improve vascular function in healthy humans

In our study peripheral blood pressure and CAP decreased, whereas FMD, as a marker for endothelial function, increased in the CF group. These findings corroborate our previous findings that not only in diseased but also in healthy individuals, CFs improve vascular function and cardiovascular risk markers (15, 40, 41). The proposed increase in eNOS activity in healthy adults (42) is one likely mechanism by which CFs and especially (-)-epicatechin improve vascular function, blood pressure (15, 23) and endothelial integrity (43). Overall dietary interventions have been shown to reestablish disturbed redox signaling in the vasculature (44). As already demonstrated in the COSMOS trial (45) a 27% lowering in cardiovascular deaths can be achieved through CF intake, at the same daily amount as given in the current study, in elderly individuals free from CVD after a 3.6-year follow-up. The amount of dietary CFs given in our study is above the average intake in a typical European diet. Dietary extracts of CFs are generally considered save in concentrations up to 1g/day and beneficial in the prevention of CV death (45). These results serve for indirect validation of the CF effect. Importantly, we saw no non-responder regarding CF effect on FMD in the intervention group.

## Data Availability

All data produced in the present work are contained in the manuscript.

https://www.example.com

## Limitations

A few limitations of the present study are worth mentioning. First, we did not assess individual food diaries addressing estimates of habitual CF intake in the participants’ regular diet. Second, we did not collect plasma nor urine levels of circulating flavanols and their structurally related or gut microbiome derived metabolites to ensure regular intake and sufficient absorption. However, the observed selective increase in FMD all participants within the CF group with a 10% percent response rate argues for a high adherence within the entire study cohort.

## Conclusion

We here provide evidence, that intake of CFs improve markers of cardiorespiratory fitness in healthy elderly humans highlighting their potential for supporting maintenance of cardiovascular health and fitness with increasing age. Future studies should focus on the evaluation, if these effects are sustained when CF are consumed in the long term and translate into sustained and relevant improvements of fitness, quality of life and lowering of cardiovascular events and mortality.

## Abbreviations

BMI: body mass index
BP: Blood pressure
BSA: body surface area
CAP: central aortic pressure
CF: Cocoa flavanol
CPET: cardiopulmonary exercise testing
CVD: cardiovascular disease
FMD: flow-mediated vasodilation
HR: heart rate
PeakVO_2_: maximum oxygen consumption
RA: radial artery
RER: respiratory exchange ratio
VO_2_: volume flow of oxygen
VT: ventilatory threshold

## Acknowledgment

Cocoa flavanol and placebo capsules were donated by Mars, Inc., USA.

## Contributions

M.G., R.S., C.H., M.K. and R.E. designed the study; M.G., N.O. and N.K. conducted the research; M.G., N.O., N.K., B.S. and R.E. analyzed the data; M.G., D.D., C.H., C.J., M.K. and R.E. wrote the manuscript.

## Graphical Abstract

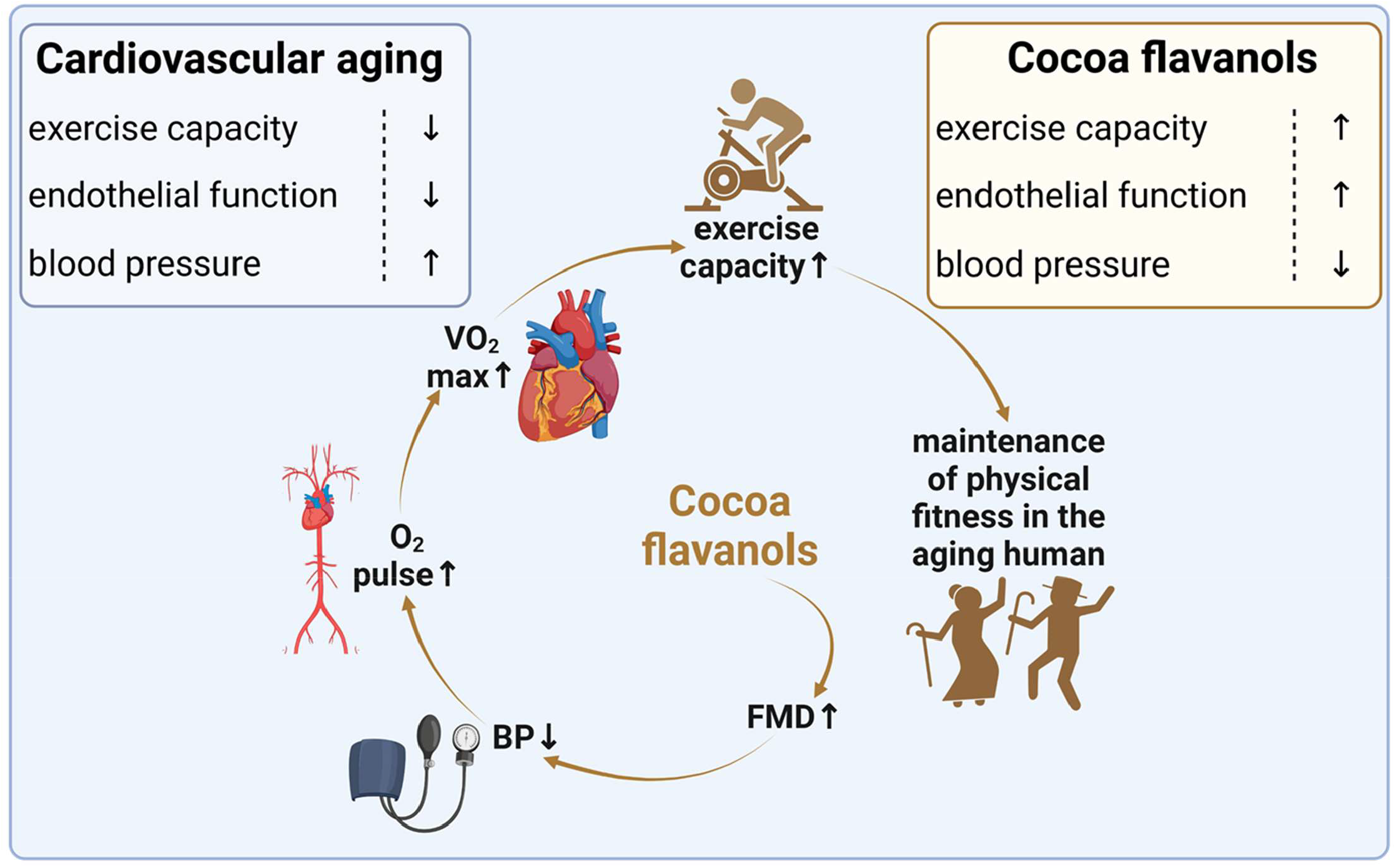

## Supplementary information

**Table S1:**
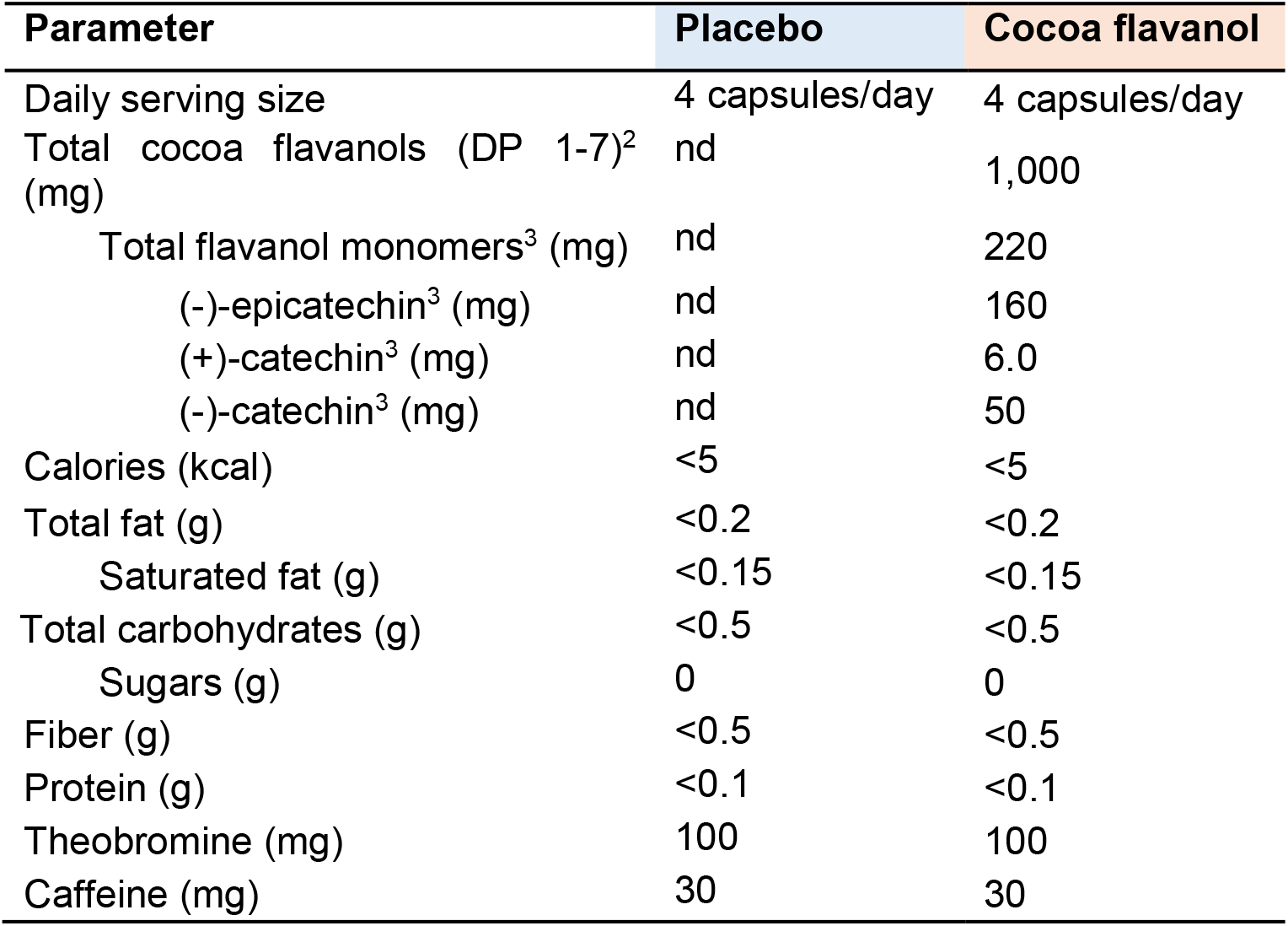
Composition of daily cocoa flavanol and placebo interventions. The participants consumed 2 of capsules in the morning with breakfast and 2 at night with dinner. nd = not detected. ^1^ Test material is not a significant source (≤1 mg/serving) of sodium, potassium, iron, magnesium, copper, manganese, phosphorous, or calcium. ^2^ Analysis based on AOAC 2020.05. Cocoa flavanols includes flavanol monomers and procyanidins with a degree of polymerization (DP) up to 7 units. ^3^ Analysis based on AOAC official method. AOAC 2020.05-2020 Flavanol and Procyanidin (by Degree of Polymerization). Available from: http://www.aoacofficialmethod.org/index.php?main_page=product_info&products_id=3046

